# First indication of the effect of COVID-19 vaccinations on the course of the outbreak in Israel

**DOI:** 10.1101/2021.02.02.21250630

**Authors:** H. De-Leon, R. Calderon-Margalit, F. Pederiva, Y. Ashkenazy, D. Gazit

**Affiliations:** INFN-TIFPA Trento Institute of Fundamental Physics and Applications, Via Sommarive, 14, 38123 Povo TN, Italy; European Centre for Theoretical Studies in Nuclear Physics and Related Areas (ECT*), Strada delle Tabarelle 286, I-38123 Villazzano (TN), Italy; Hebrew University-Hadassah Braun School of Public Health, 9112102, Jerusalem, Israel; Dipartimento di Fisica, University of Trento, via Sommarive 14, I–38123, Povo, Trento, Italy; Racah Institute of Physics, The Hebrew University of Jerusalem, 9190401 Jerusalem, Israel

## Abstract

Concomitantly with rolling out its rapid COVID-19 vaccine program, Israel is experiencing its third, and so far largest, surge in morbidity. We aimed to estimate whether the high vaccine coverage among individuals aged over 60 years old creates an observable change in disease dynamics. Using observed and simulated data, we suggest that the shape of the outbreak as measured by daily new moderate and severe cases, and in particular of patients aged over 60, has changed because of vaccination, bringing the decline in new moderate and severe cases earlier than expected, by about a week. Our analyses is consistent with the assumption that vaccination lead to higher than 50% protection in preventing clinical disease and with at least some effectiveness in blocking transmission of elderly population, and supports the importance of prioritizing vulnerable population. This is the first indication of the effectivity of COVID-19 vaccine in changing the course of an ongoing pandemic outbreak.

**One Sentence Summary:** We show, by data analysis and modelling of the dynamics of COVID-19 pandemic in Israel, that the current nationwide outbreak, that had up to 0.1% of the population confirmed daily, is clearly affected by the vaccination program, that reached a coverage of more than 80% among people ≥ 60 years old.

## Main Text

The development of SARS-COV-2 vaccines provide hope for the culmination of the global pandemic and return to normal life. So far, the FDA gave emergency authorization approvals (EUA) to two mRNA vaccines – the Pfizer-BioNTech and Moderna vaccines. Both have shown similar overall efficacy of 94-95% in preventing clinical disease in phase three trials and received emergency use approvals from the FDA for use in individuals aged 16+ and 18+ years, respectively (1, 2). Similar efficacy was found for prevention of severe disease, although numbers were small (overall, 9 and 30 severe cases in the Pfizer and Moderna trials, respectively). It is yet unknown whether these vaccines are effective in preventing transmission, although preliminary evidence from the Moderna trial suggest that there may be some level of reduced transmission following immunization (2). Furthermore, the confidence intervals of the efficacy estimates in participants aged 65 years or more are wide and call for further studies.

Israel has experienced several distinct surges of morbidity, leading to three national lockdowns, the latest being imposed on January 8^th^, 2021. Following the FDA EUA approvals, Israel started its vaccination program on December 19^th^, 2020 (3), using almost exclusively the Pfizer-BioNTech vaccine and prioritizing individuals aged 60+ years, immune deficient patients, and healthcare workers to be the first vaccinated. Since then, and up to date (January 25, 2021), about 2.6 million Israelis (29% of the population) have been vaccinated with at least the first dose, and the second dose is rolling out, yielding vaccine coverage of 80% in the ≥60 years age group, as published by the Israeli Ministry of Health (4). Thus, monitoring epidemic evolution in Israel may allow assessment of the effectiveness of vaccines in mitigating the outbreak on a national level. A major challenge in this respect is to identify a distinct indicator of the effect of vaccines differentiating it from the effect of lockdown.

We used data analysis tools and simulations to model epidemic dynamics in Israel, comparing different scenarios of lockdown duration and effectiveness. Each scenario is modeled with and without the up-to-date vaccine coverage. We studied the daily trends of new confirmed cases as well as the trends of new moderate or severe COVID-19 cases in Israel. Moderate COVID-19 is defined as a case positive for SARS-COV-2 in RT-PCR test, with bilateral pneumonia. A severe case of COVID-19 is defined in Israel as a SARS-COV-2 confirmed case with oxygen saturation below 93% while breathing ambient air (these include also critical cases). Therefore, as long as hospitals are not overwhelmed, numbers of moderate and severe cases are independent of either testing policy or availability. In addition, these cases are at increased risk for further deterioration, with 15-22% fatality rate within 10 days (See Methods Section B). On average, moderate and severe admissions take place about 5 days following a positive RT-PCR test, and an estimated 10 days from the actual infection.

Thus, the number of new moderate and severe cases makes a clear and robust indication for the dynamics of disease, that has the advantage of being evident quite soon after infection. Finally, elderly individuals are at increased risk for having moderate or severe COVID-19. Therefore, we studied the trends of moderate and severe cases as candidate measures of disease dynamics and the effect of vaccine on the outbreak.

We start by analyzing the status of the pandemic in Israel, using the effective reproduction number, *R*_*e*_, which is approximated using the 4-day growth factor of the weekly average of new COVID-19 cases (5). In Fig-1(a) we show that the dynamics leading to the January lockdown, as described by changes in *R*_*e*_ are quantitatively similar to those leading to the second lockdown in Israel, which was implemented on September 19, 2020. The effective *R*_*e*_, based on the daily admissions of moderate or severe cases shifted by five days (in agreement with clinical course), are highly correlated with the effective *R*_*e*_ based on confirmed cases and show similar correlation with the reproductive numbers shown in September (Fig S-2). The agreement of reproductive numbers based on different measures supports the notion of similar dynamics in both waves, which is not surprising considering the similarities in the non-pharmaceutical intervention restrictions during the lockdowns. It also minimizes the concerns of systematic errors, such as under-detection of cases. At the onset of both nationwide lockdowns, the effective *R* ranged 1.1-1.3.

**Figure 1-(a):**
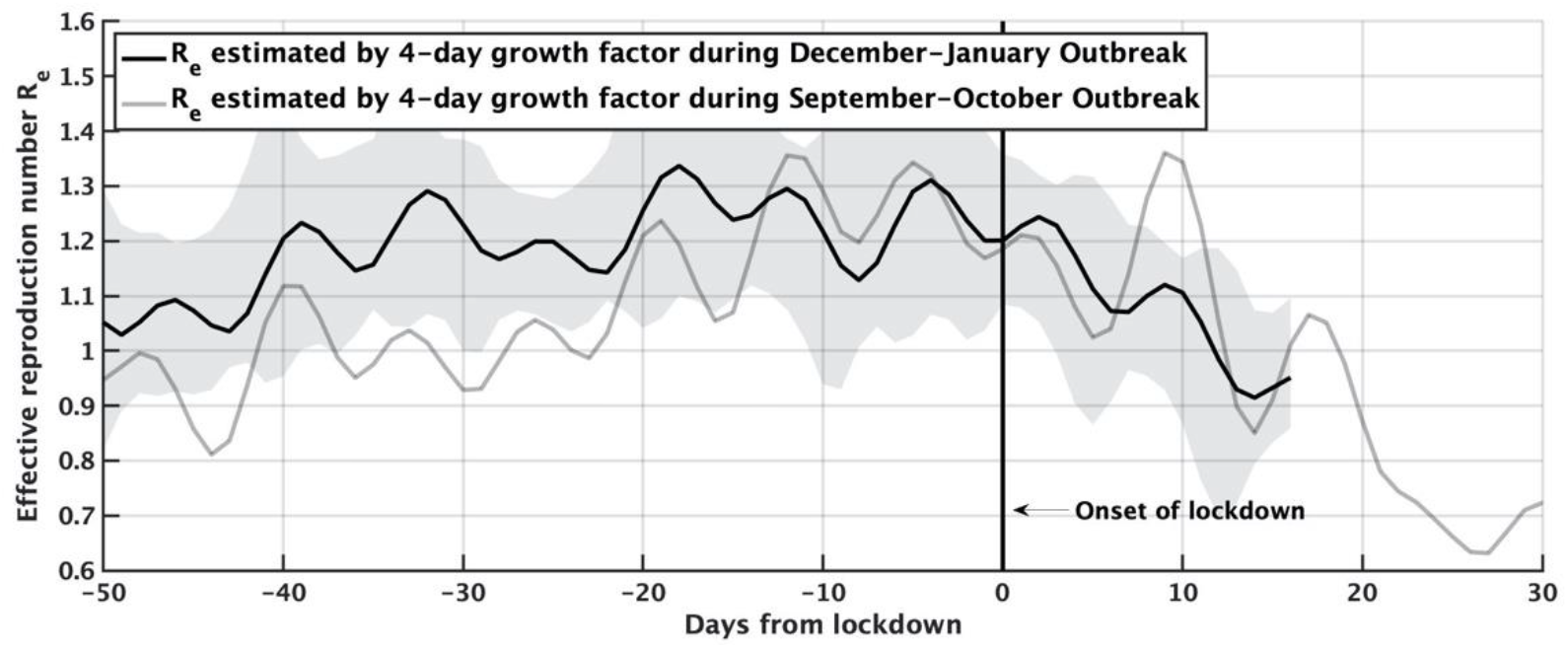
Effective reproduction number R_e_ in Israel, as estimated by the 4-day growth factor of daily confirmed cases, with respect to the onset of the national lockdowns either on Sep 19, 2020 (gray line), or Jan. 8, 2021 (black line). Shaded area denotes 1.5 standard deviations about R_e_ of the black line. One observes the striking similarity between R_e_’s, which represents the dynamics of the outbreak, between the two lockdowns.

This similarity is observed also in Fig-1(b), where the absolute number of the 7-day moving average of the daily-confirmed cases, shows the same dynamics starting about three weeks prior to lockdowns, albeit the third wave is about 35% larger, due to late entrance to the January lockdown.

**Figure 1-(b):**
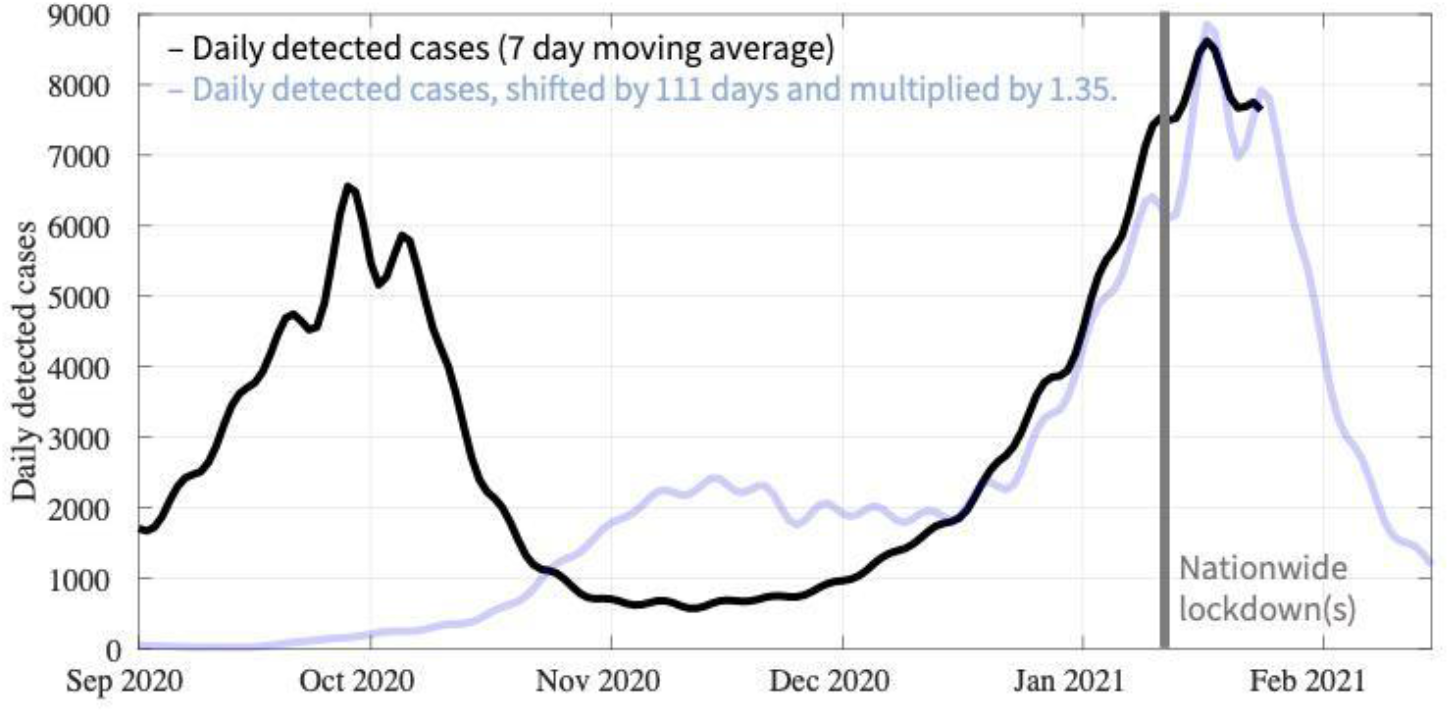
Daily detected cases (7-day average) in Israel, since September. The light blue line is the black line shifted by 110 days (so that the onset of lockdowns coincides) and multiplied by 1.35 to match for the increased size of the third wave. One sees that while the evolution of the outbreaks is extremely similar, the number of cases in January is bigger by 35%, than the corresponding date in September.

Fig-2 shows that similarities between the second and third lockdowns discontinue. While the number of new moderate and severe cases among patients older than 60 has peaked on the 6^th^ day of the third lockdown and decreased thereafter, in the second lockdown it reached its peak on the 14^th^ day of the second lockdown, and decreased thereafter. Such a time difference is not found when studying the daily admissions of moderate or severe cases among younger patients. In fact, as of today (January 25, 2021), the current outbreak has not seen a decrease in the latter daily number, as can be seen in Fig. 3-d.

**Figure 2:**
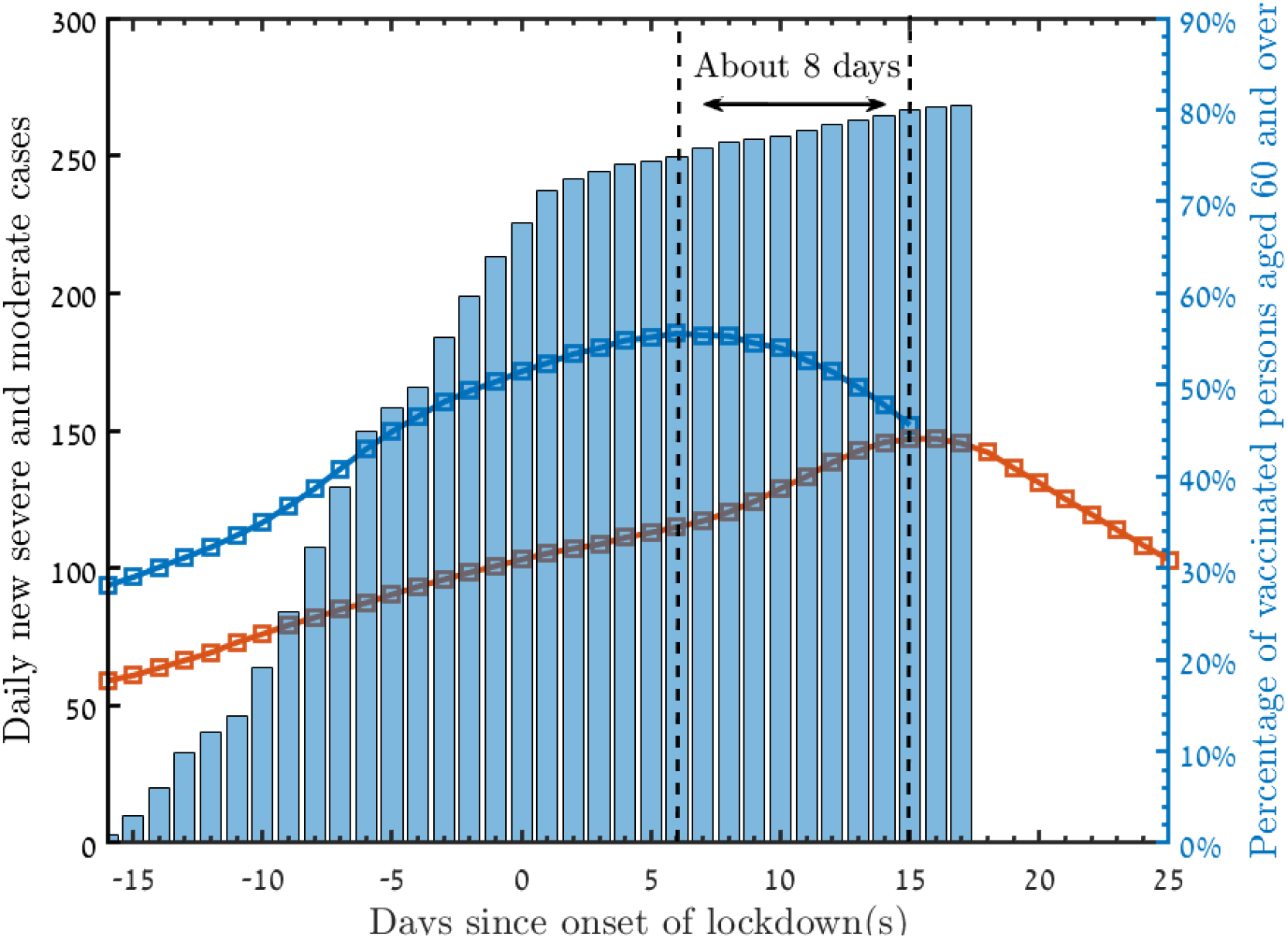
New daily moderate and severe cases among patients older than 60 in the January lockdown (blue), and September lockdown (red). The bars represent the percentage of vaccinated people older than 60 years old, relative to the January lockdown. The time difference of about 8 days between the peaks is an indication for the vaccination effectiveness.

**Figure 3:**
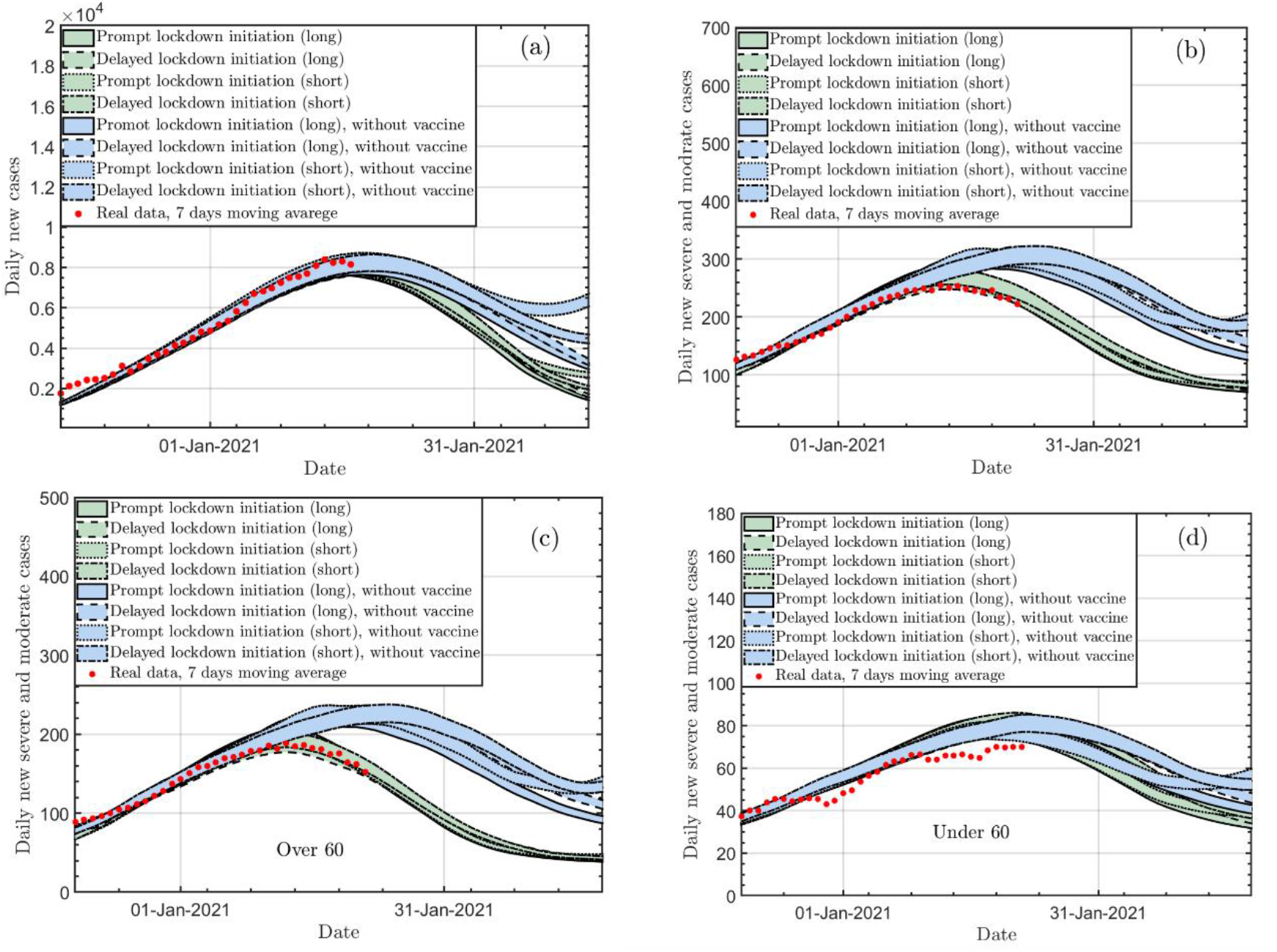
Expected morbidity according to different scenarios of lockdown duration and strictness, and according to vaccinations, following the rollout in Israel. a. Daily new confirmed cases b. moderate and severe cases c. moderate and severe cases among individuals aged 60+ years d. moderate and severe cases among individuals younger than 60 years. For all panels: Green (blue) solid line: long prompt lockdown with (without) vaccinations; Green (blue) dashed line: long delayed lockdown with (without) vaccinations; Green (blue) dotted line: short prompt lockdown with (without) vaccinations; Green (blue) dashed-dotted line: short delayed lockdown with (without) vaccinations.

We hypothesize that having an early peak and a decline is a result of vaccinations, and suggest the time difference between peaks with and without vaccination as an indicator for vaccination effectiveness. In order to test this hypothesis, we employed homogenous random movement models as introduced by De-Leon and Pederiva (6) to study the expected effects of vaccinations on the dynamics of the moderate and severe hospitalizations during the January lockdown. The models take as input the effective repruduction number as estimated by the detected cases, and use them to model the dynamics of infections, and the resulting moderate and severe cases.

Since the estimates of the infection level include some uncertainties, e.g., due to testing policies, we use various scenarios to estimate the lockdown effect, ranging from 2-5 weeks of length, and assuming either prompt or delayed compliance of the population leading to prompt or delayed effect on *R*_*e*_. We additionally assume that the third lockdown yields similar effects as second lockdown, i.e., reaching *R*_*e*_ of 0.7 (see Fig.1-a). The details of the scenarios are given in Section A of the Methods. We employed models with and without vaccines; the models with vaccines assume coverage according to vaccine rollout in Israel. The effectiveness of vaccines in preventing symptomatic disease was taken as 0% in the first week after the first dose, 30% on days 7-10, 60% on days 11-20 and 95% from day 21 (second dose) onwards. We first assume that vaccinations prevent infections, though we check the sensitivity of the results to this assumption later on, as this was not reported in the Pfizer-BioNTech trial.

Figure 3 presents the results of these simulations. In general, one observes that the effect of the properties of the lockdown are not important for the dynamics in January, in particular for the simulations that include vaccinations. The predicted daily confirmed cases in the different scenarios, as shown in Fig.3-a, are expected to peak on similar timing in all scenarios, though the rate of the following decline changes according to the scenario, and more importantly, according to whether vaccines were administered, with a steeper decline when the population is vaccinated. This is expected, under the assumption that vaccinations prevent transmission.

An important observation appears when studying the predicted numbers of moderate and severe cases according to these scenarios. In Fig. 3-b, a time difference of 6-11 days is predicted between the peaks of new daily moderate and severe cases of vaccinated and unvaccinated scenarios. Stratifying the models by age (Fig 3-c and 3-d), it is evident that this bringing forward of the decline in moderate and severe cases is limited to patients older than 60, while it is not expected to affect the timing of peak and decline in those younger than 60.

As aforementioned, there are still open questions regarding the effectiveness of vaccines, in particular, whether the 95% efficacy achieved in clinical trials translates to similar effectiveness in preventing clinical disease in a national vaccination campaign. In addition, the effectiveness in preventing transmission is unknown. We therefore employed simulation models with sensitivity analyses (Fig 4). Trivially, the confirmed cases are not expected to be affected by vaccinations that do not prevent infection (fig.4-a). However, we see that the indicator we suggested here, i.e., the time difference in the peaks in the new daily moderate and severe cases between vaccinated and unvaccinated scenarios is shortened by about 1-2 days, if the effectiveness of the vaccinations is 95%, even if transmission is not prevented by the vaccines. In addition, we find that the indicator changes by about a day if the 95% effectiveness is reached only on day 28 after the first dose.

**Figure 4:**
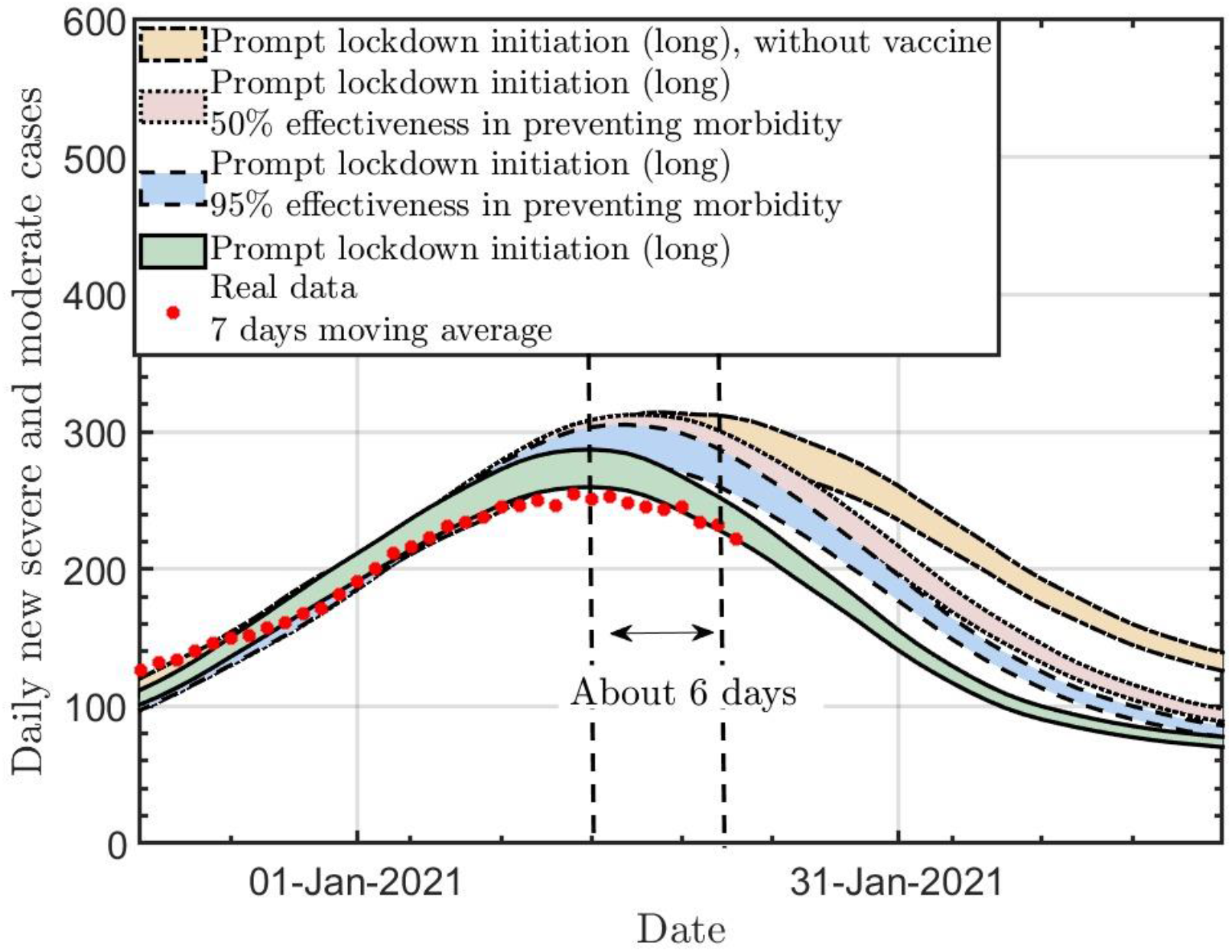
The new daily moderate and severe cases for three different vaccine effectiveness scenarios, and without vaccines. Solid line: Long effective lockdown with an effective vaccine. Dashed (dotted) line: Long effective lockdown with 95% (50%) effectiveness in preventing morbidity, while not preventing infections.

The indicator loses its significance, when assuming vaccine effectiveness decrease to 50% in the latter scenario.

Summarizing, in this short term estimation of the effects of vaccines on the dynamics of Israel’s current outbreak, we suggest the time difference between the observed dynamic of the new moderate and severe COVID-19 cases under a vaccination rollout to the expected dynamics, comparing to a previous wave when no vaccines were available. Both observed data and models suggest that indeed, prioritizing to vaccinate the elderly was effective in reducing the influx of moderate and severe cases, and bringing the tipping point earlier than expected, by about a week, has there not been any vaccines. Since length of stay is relatively long (10-20 days), the change in admitted patients is not detected immediately in the total numbers of hospitalized patients.

Previous modeling of vaccines looked into a possible impact of vaccines on the outbreak in the US. According to this study, vaccine coverage that will reach 40% in the American population within nearly a year will mitigate morbidity effectively, even though this is far from reaching herd immunity (7). Nevertheless, the modeling ignores current trends in the dynamic of COVID-19 in the US, except for the cumulative incidence of the disease so far. Moreover, this study assumes that vaccination completely blocks transmission, and that asymptomatic disease is not associated with transmission at all. Another modelling study that compared different strategies in prioritizing vaccines (8), suggested that prioritizing individuals aged 60+ years (direct protection) is more effective when transmission is high (*R*_*e*_ ≥ 1.5), whereas prioritizing younger individuals is more effective when transmission is low (*R*_*e*_ ≤ 1.2). In all scenarios they assumed rollout rate of 0.1-0.76%. In Israel, those aged 60+ were prioritized in the first month of vaccines and are now gradually being offered to younger individuals. The rollout rate achieved in some days 2% of the population (3).

Our study has some limitations. First, our models assume that the third lockdown would have similar effectiveness in curbing the third wave as the second lockdown if no vaccines were employed. It should be noted that previous lockdowns took place around holidays where people usually work less. Whether because of no holiday, “pandemic fatigue”, or loss of trust in the government or healthcare systems (9, 10), it is quite possible that the public adherence to the current lockdown is reduced, impairing its effectiveness. Second, our models assume equal chance encounters between individuals. We could not model social interactions, as there is no such data available for Israel. Israel is a country with the highest fertility rate in the OECD countries (11) which translates into big households and crowded classrooms and different social interactions than those observed in European or North American countries. This minimized the possibility to adopt social connections surveys conducted in other countries. Moreover, like any other country, there are sectors in the population that co-interact within the sector than with members of other sectors. This was not taken into account, based on the notion that outbreaks that started within specific population sects (e.g., Israeli Arabs, ultra-orthodox Jews) expanded to the general public and to national outbreaks, leading both to the second and third waves.

To the best of our knowledge, this is the first study to estimate the effect of mass vaccinations in a tome of a pandemic using a new indicator. While our study cannot accurately estimate the effectiveness of the Pfizer-Biontech vaccine, it suggests that the effectiveness is greater than 50% and that there is a considerable level of prevention of transmission following vaccinations.

## Data Availability

The data that support the findings of this study are available from the corresponding author upon reasonable request:
* Most of the data is available on https://data.gov.il/dataset/covid-19
* Daily moderate and severe cases by age groups were obtained from the Ministry of Health. Data provided by the Ministry of Health was available through the Ministry's research room, accessible only to authorized researchers (Ronit Calderon-Margalit). Data was then aggregated by age and analyzed in two age groups and without any identifiers.

https://t.me/MOHreport/7485

https://www.rki.de/DE/Content/Infekt/EpidBull/Archiv/2020/Ausgaben/17_20.pdf%20?__blob=publicationFile

https://data.gov.il/dataset/covid-19

## Acknowledgements

DG, RCM and YA gratefully acknowledge the discussions with the members of the Hebrew University COVID-19 Monitoring Team, M. Assaf, N. Katz, R. Nir-Paz, and M. Bing. DG and HDL thank Nadav Eyal and Eldad Sitbon for fruitful discussions.

## Notes

### Competing Interest Statement

The authors have declared no competing interest.

### Funding Statement

No external funding was received

### Author Declarations

The study protocol was approved by the Institutional Review Board (Helsinki Committee) of Hadassah Medical Center. Study was exempted from informed consent based on anonymity of participants (anonymization carried out by the Ministry of Health) and aggregated data analysis.

