## Supplementary material for "First indication of the effect of COVID-19 vaccinations on the course of the outbreak in Israel": Methods

**Supplementary materials**

**Methods:**

**A – The model**

The simulations in this work use previously described models (6). Briefly, the models simulate the population as a set of interacting individuals, employing the basic mechanism of statistical physics. Each entity is tagged with health status (susceptible, infected, recovered/dead or vaccinated). The models use standard thermodynamical parameters (temperature and density) to describe mixing and the number of average encounters of individuals. Temperature, in statistical mechanics, is the average kinetic energy, and it is directly related to diffusion and mobility. which in the current case is reciprocal to the average time and spatial rate of population mixing. Density is a measure of the average distance between individuals. Meetings between individuals lead to transitions between health states, using clinical infection probabilities, based on the most recent epidemiological data. The models can be applied to different scenarios, e.g., a city suburb population, and a single university classroom. The algorithm is based on standard Monte Carlo (MC) procedures of sampling the transition among subsequent states, which are essentially sampled from a statistical distribution (12).

The model is a "one-way" Ising-model Monte-Carlo simulation, in which a healthy, susceptible, person (i) can be infected with a daily probability, $P_i = \sum_j P_{ij}$ , where $P_{ij}$ is a function of the distance, $R_{ij} = |\vec{\boldsymbol{r_i}} - \vec{\boldsymbol{r_j}}|$, between each infected person (j) in the surface and the healthy person (i). Based on the epidemiological data (12), we assume that an infected individual can infect susceptible people in its environment from the 3rd day of infection to the 7th day. A "vaccination" compartment which switches a vaccinated person from the susceptible state to the immune state (see Ref.(6) for more details). The infection probability, $P_i$ of a person (i) given the surrounding is given by $\sum_{\boldsymbol{j=1}}^{\boldsymbol{n_{sick}}} \boldsymbol{exp}\left[-\frac{(|\vec{r_i}-\vec{r_j}|)^2}{2\cdot\sigma_r^2}\right] \times \boldsymbol{f}$, with $\boldsymbol{n_{sick}}$ the number of sick entities/individuals, $\sigma_r \approx 2.4m$ is the physical typical range of infection, and $\boldsymbol{f}$ is a factor which depends upon behavioral characteristics, e.g., number of times a person leaves home every day, household infection characteristics, etc., as well as whether the (j) person shows symptoms.

In the Monte-Carlo application, one then adds a uniformly distributed random variable with a [0,1] range to this probability. The person is infected if the total result is greater than 1. In the current work the simulation is calibrated to reproduce given daily effective reproduction number $R_e$ starting from a given initial daily confirmed case. The change in $R_e$ is modeled by a change in the density of the system.

Thus, the important parameters are just the transition probabilities, vaccine efficiency and rate of population vaccination (6), as well as the $R_e$ history. The $R_e$ that is used as an input to the simulation is the weekly averaged 4-day growth factor in Israel, until the onset of the third lockdown (January 8, 2021).

Figure S-1 shows the agreement of the simulated daily new cases to the data in Israel from August 1, 2020 till January 6, 2021 based on the effective reproduction number, $R_e$ in Israel (5). This bolsters the validity of the model and enables us to use it in this work for future predictions.

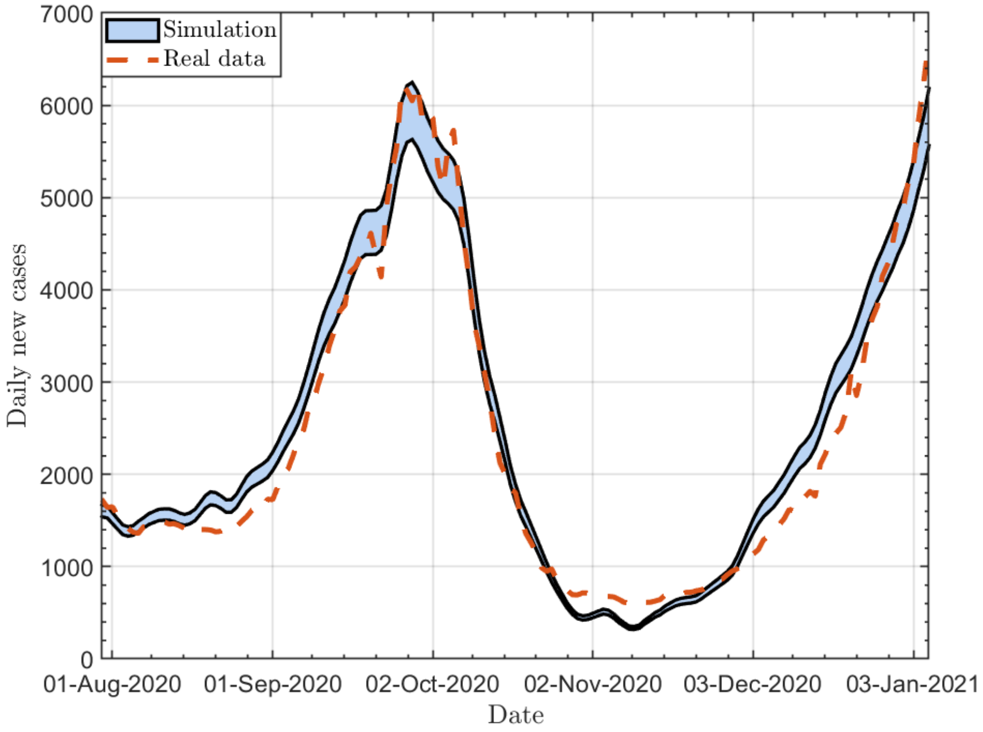


*Figure S1: The daily new confirmed cases between Aug 1st and Jan 6th, 2021. Solid line: simulation. Dashed line: real data.*

From January 8, 2021, when the government imposed a strict lockdown, population compliance is simulated in different ways. Prompt compliance implies a reduction in infections immediately as the government ordered lockdown, while a delayed compliance implies such reduction a week after the government ordered lockdown. The reduction in infections has a delayed manifestation, since individuals are being tested either when they develop symptoms or when they are aware of being a contact of a confirmed case. Thus we assume that prompt (delayed) compliance means that $R_e$ reduces gradually, starting January 12th (19th), 2021, to the typical effective reproduction number in lockdowns in Israel of 0.7 ($R_e$=0.7). The lockdown usually takes place for 2-5 weeks, followed by reopening which is modelled as exponential increase towards $R_e$=1.2-2. The reopening is beyond the scope of this paper, as it does not affect the indicator we introduced in measuring the short-term effect of vaccination.

The different scenarios used for the simulated scenarios are depicted in Fig. S2.

Infected cases could deteriorate into moderate or severe cases within 5 days from positive test results (as explained in Section B below). We use fitted data of Israeli hospitalizations to model this by age. The fit appears in Section C.

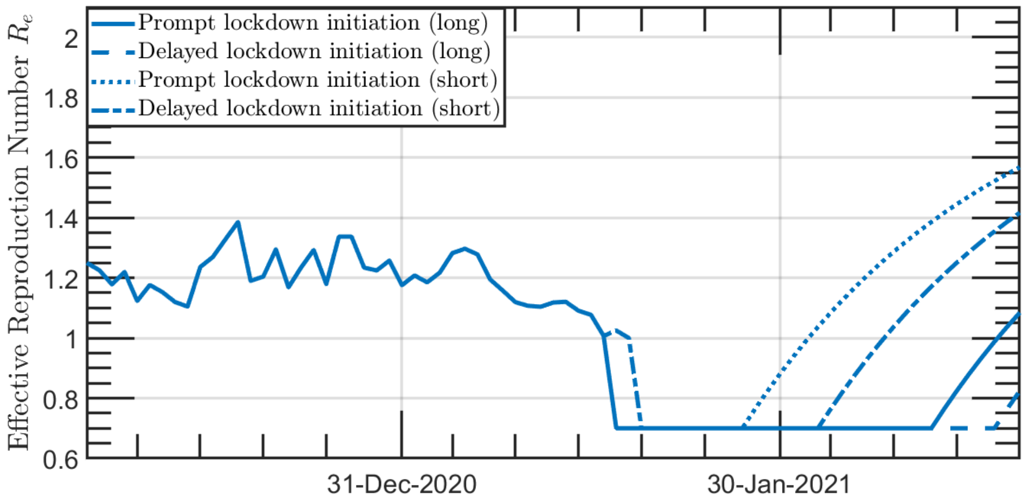


*Figure S2: The confirmed cases effective reproduction number of the virus, $R_e$, as a function of time for four different scenarios. Solid line: four-weeks prompt lockdown staring at Jan 12$^{th}$ (for confirmed cases). Dashed line: four-weeks delayed lockdown staring at Jan 19$^{th}$. (for confirmed cases). Dotted line: two-weeks prompt lockdown staring at Jan 12$^{th}$ (for confirmed cases). Dotted-dashed line: two-weeks delayed lockdown staring at Jan 19$^{th}$ (for confirmed cases).*

### B – Moderate and severe daily admissions as an indicator for the state of the pandemic in Israel

In this work, we use moderate and severe daily admissions as a measure for the disease in Israel. This is justified by the fact that there exists a correlation between this parameter and the population infection rates. This is exemplified in Fig. S3, which showed that there is an agreement between the effective reproduction number estimated by the 4-day growth factor of either detected cases (full line), with its standard deviation (shaded area), or using the moderate and severe new daily hospitalizations (dashed line). Maximal correlation is achieved by shifting back the $R_e$ of the new detected cases by 5 days. As confirmed cases are delayed by 5 days on average from infection in Israel, this is consistent with a 10-day average deterioration time from infection to moderate or severe condition.

In addition, we find that fatalities are also strongly correlated to moderate and severe new daily hospitalizations. In fact, the number of daily fatalities in Israel on a given day is given by 15-22% of the new cases hospitalized exactly 10 days ago. (Fig. S4)

As a result, this parameter is a clear and robust indicator for a COVID-19 serious case, that has the advantage of being evident quite promptly after infection, compared to death, but does not depend on detection or population test compliance. Thus, it is ideal as an objective measure of disease dynamics and the effect of vaccine on the outbreak.

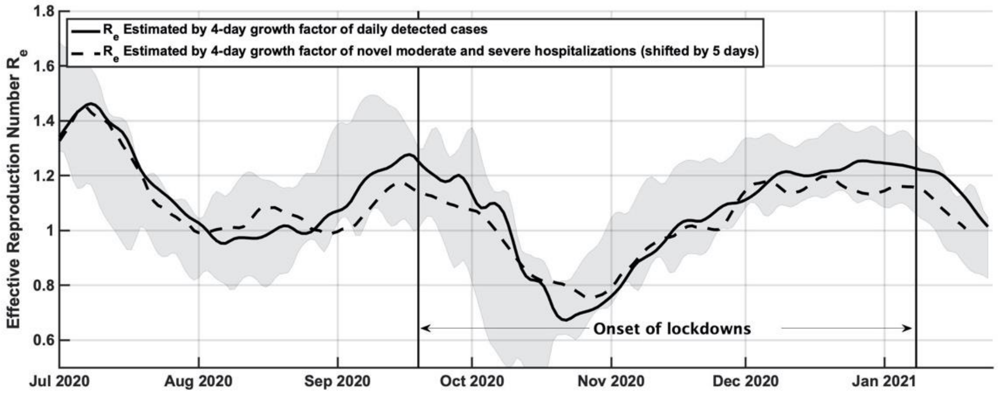


*Figure S3: Effective reproduction number estimated by the 4-day growth factor of either detected cases (full line), with its standard deviation (shaded area), or using the moderate and severe new daily hospitalizations (dashed line), shifted back 5 days to maximally correlate to the detected cases.*

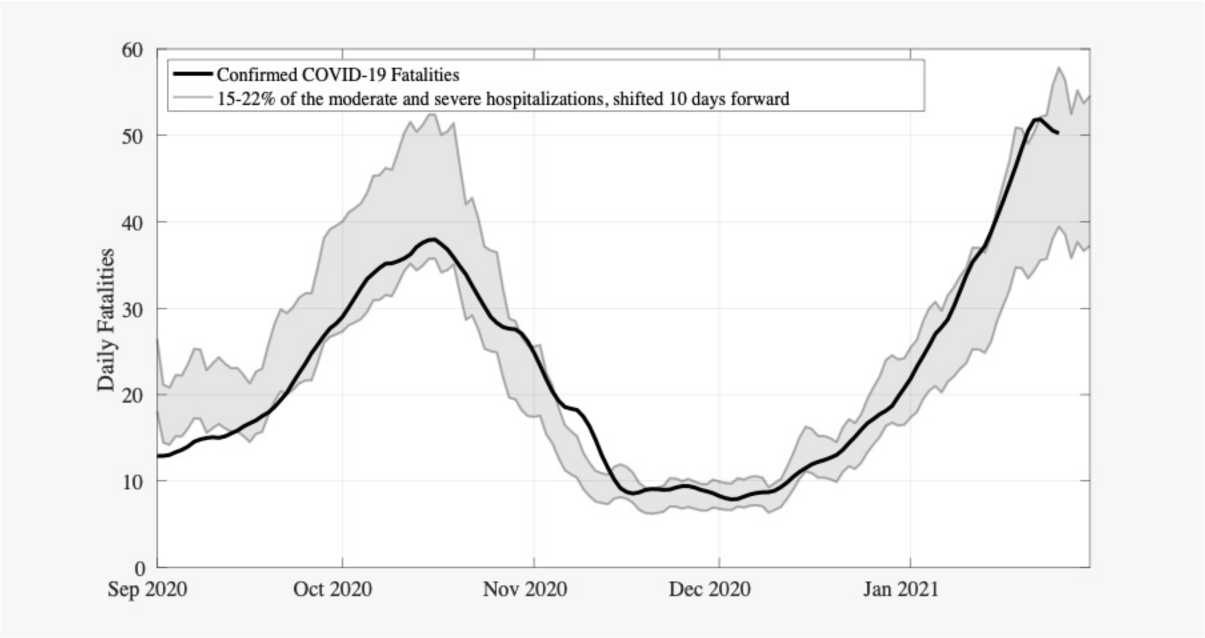


*Figure S4: daily fatalities (line), and a model for the fatalities assuming that 15-22% of new moderate or severe cases die within 10 days.*

### C – Simulating the time dependence of new moderate and severe cases

The input $R_e$ allows a prediction of the evolution of confirmed cases. As shown by using Fig. S3, this leads to admissions in moderate or severe condition. We then calibrated the simulation to reproduce the new moderate and severe cases around the second lockdown period (i.e., between Aug 1$^{st}$, 2020 and Oct 31$^{st}$, 2020). The calibration process leads to a fit of the probability of a person to be hospitalized in this condition. It is found that the probability of hospitalization decreases as the load on hospitals increases. This is a measure of the effect of hospital capacity on the treatment an individual receives.

Fig. S5 presents the percentage of moderate and severe morbidity as function of the daily confirmed cases (with a five-day shift) for patients aged over and under 60 years. Data and their fitted analytical functions are presented for elderly (over 60) and younger (under 60) patients, the severe/moderate morbidity in the current wave can be estimated for both age groups. These functions are then used to predict such morbidity.

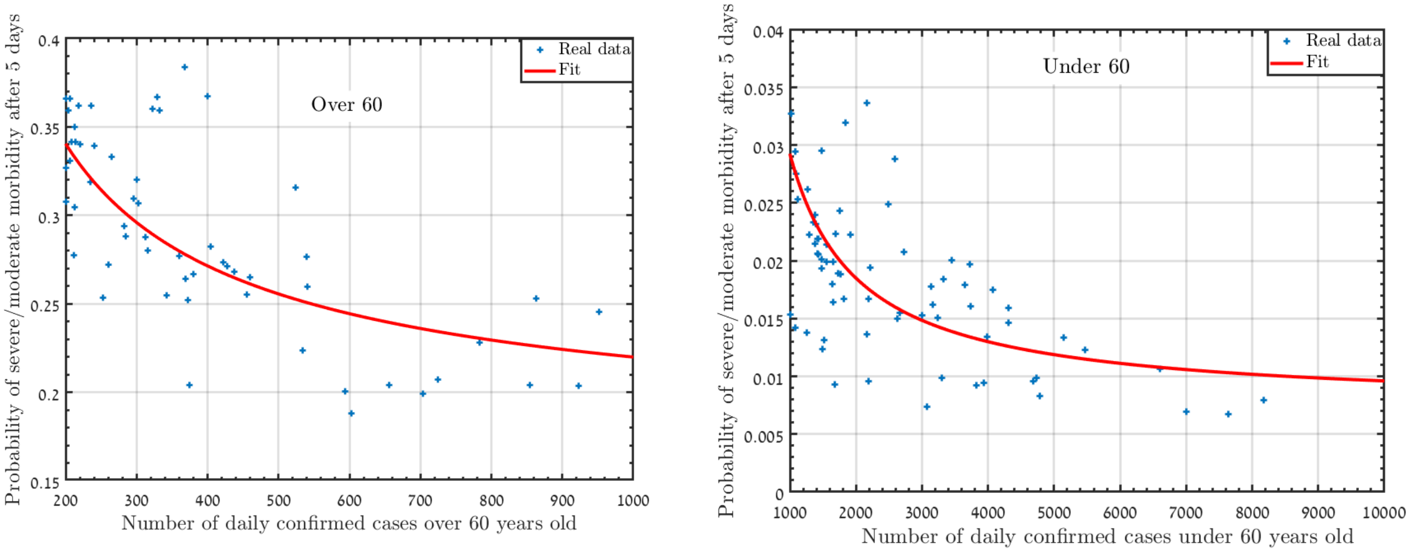


*Figure S5: the probability of moderate and severe morbidity as a function of the daily confirmed cases (with a five-day shift) for patients aged over (left panel) and under (right panel) 60 years old.*

*For both panels, the dots represent real data of the moderate and severe morbidity in Israel between 01/08/2020 and 31/10/2020. The solid line is fitted function.*
